# Erythrogram directly from the microscope eyepiece: a feasibility study using artificial intelligence

**DOI:** 10.64898/2025.12.23.25342416

**Authors:** Lucas Praciano

## Abstract

**Background:** Erythrocyte indices are essential for the diagnosis and monitoring of hematologic diseases, but their determination depends on automated hematology analyzers, which limits access in regions with limited laboratory infrastructure. Although artificial intelligence approaches have been proposed for hematologic analysis, they usually rely on slide scanners or digitization systems. To date, no validated approaches have been identified in the literature that estimate these indices directly from images obtained through the eyepiece of conventional optical microscopes.

**Objective:** To evaluate the feasibility of automated prediction of erythrocyte indices from blood smear images obtained directly through the eyepiece of conventional microscopes using convolutional neural networks.

**Methods:** Two hundred blood samples stained using the May-Grünwald-Giemsa method were analyzed and photographed using a standard optical microscope. Four architectures, DenseNet-121, EfficientNet-B0, ResNet-18, and ResNet-34, were evaluated at different resolutions using 10-fold K-Fold cross-validation.

**Results:** For RBC, HGB, and HCT, ResNet-34 at a resolution of 1024*×*1024 pixels achieved superior performance, with *R*^2^ between 0.90 and 0.92, Pearson correlation *r >* 0.95, and mean absolute errors of 0.184 *×*10^6^*/µ*L, 0.524 g/dL and 1.292%, respectively. For RDW-CV, DenseNet-121 achieved *R*^2^ = 0.49 and *r* = 0.71, reflecting the greater complexity of this parameter. Bland–Altman analysis confirmed adequate agreement, with biases close to zero and more than 94% of observations within the limits of agreement.

**Conclusion:** Artificial intelligence demonstrated excellent predictive performance in estimating the erythrocyte indices RBC, HGB, and HCT, with *R*^2^ *>* 0.90, from images obtained using a conventional microscope and accessible hardware. This approach has significant potential to democratize access to hematologic analysis in resource-limited settings, although multicenter validation and regulatory evaluation are required before clinical implementation.

## 1 Introduction

Among the components of the complete blood count, the erythrogram describes red blood cell parameters through measures such as red blood cell count, RBC, hemoglobin, HGB, hematocrit, HCT, and the red cell distribution width, RDW, in addition to derived indices such as MCV, MCH, and MCHC, which are fundamental for the characterization of anemia, polycythemia, and myelodysplastic syndromes [1, 2].

Determination of these indices generally depends on automated hematology analyzers, whose availability is limited in settings with restricted laboratory infrastructure. This dependence hinders expanded access to diagnosis in small services and in resource-limited regions, motivating the adoption of alternative approaches based on more accessible equipment [3].

Artificial intelligence methods have shown consistent performance in the analysis of medical and hematologic images [4, 5]. In the specific case of blood smears, recent studies have shown that deep neural networks can predict hematologic parameters with high accuracy. However, these approaches predominantly depend on slide scanners or automated digitization systems, resulting in high implementation costs [6].

Images acquired by standard optical microscopy, especially through the eyepiece, represent a widely available and low-cost alternative, but there are still no validated approaches in the literature that directly use this type of image to estimate erythrocyte indices. Considering that parameters such as RBC, HGB, and HCT reflect global characteristics observable on the smear, such as erythrocyte density and distribution, it is hypothesized that convolutional neural networks can estimate these indices from such images.

Thus, this study evaluates the feasibility of automated prediction of erythrocyte indices using convolutional neural networks from blood smear images captured directly through the eyepiece of conventional optical microscopes.

## 2 Materials and Methods

### 2.1 Ethical Approval

The study was submitted to the Brazilian Research Ethics Committee, for review on May 5, 2025, and was approved under protocol number 88586225.9.0000.0229. The study was conducted in partnership with Laboratório Hipólito Monte, in Fortaleza, Ceará.

### 2.2 Data Collection

Residual whole-blood samples in EDTA tubes, previously processed for diagnostic purposes and intended for disposal, were used for data collection. Samples were anonymized and made available to the researcher within 24 hours after collection, preserving cell morphology.

### 2.3 Slide Preparation and Staining

Standardized blood smears were prepared using 7 µL of whole blood and the HEMAPREP device, provided by HORIBA do Brasil, which ensured procedural standardization by maintaining a constant tilt angle and smearing speed. Slides were stained using the May–Grünwald–Giemsa (MGG) method and air-dried at room temperature.

### 2.4 Microscopic Image Acquisition

Stained slides were photographed using a Digilab DI-136B binocular microscope coupled to a Wylie 48 UltraHD X60 camera with a 1/1.8 ” CMOS sensor, producing images of 3840*×*2160 pixels. A Fafeicy 0.5*×* C-mount adapter was used to couple the camera, fixed to the camera body and fitted into the eyepiece tube. Images were acquired under maximum illumination, with the condenser in the lower position, using a 4×/0.10, 160/– objective for initial localization and a 10×/0.25, 160/0.17 objective for image capture, ensuring a field of view suitable for erythrocytes. One image per sample was recorded by positioning the end of the slide tail at the center and advancing to the area of optimal cell dispersion. Figure 1 illustrates examples of the collected images.

**Figure 1:**
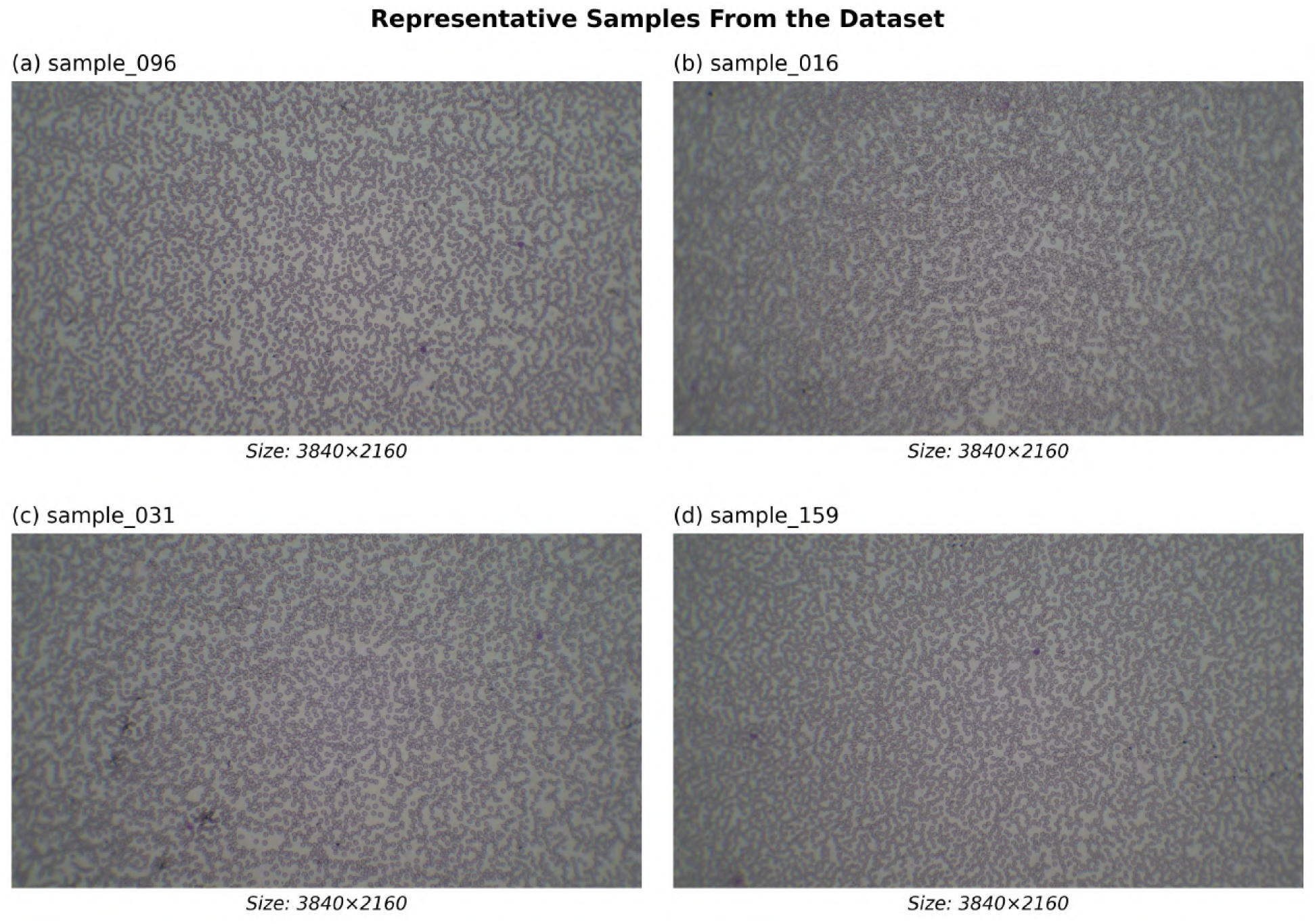
Examples of collected blood smear images. Images obtained with optical microscope at 10× objective, May-Grünwald-Giemsa stained, resolution 3840×2160 pixels. (a) Sample 096, (b) Sample 016, (c) Sample 031, (d) Sample 159, illustrating the morphological variability and erythrocyte density of the dataset.

### 2.5 Hematologic Analysis

Following slide preparation, samples were analyzed using a Sysmex XN-550 hematology analyzer. Results were recorded and subsequently reviewed by the researcher to ensure data integrity. The measurement units used in this study are described in Table 1.

**Table 1:** Measurement units adopted in this study.

| Parameter | Description | Unit of measurement |
| --- | --- | --- |
| RBC | Red blood cell count | $10^6/\mu\text{L}$ |
| HGB | Hemoglobin | g/dL |
| HCT | Hematocrit | % |
| MCV | Mean corpuscular volume | fL |
| MCH | Mean corpuscular hemoglobin | pg |
| MCHC | Mean corpuscular hemoglobin concentration | g/dL |
| RDW-CV | Red cell distribution width (coefficient of variation) | % |

### 2.6 Data Analysis

#### 2.6.1 Sample Characterization

The final dataset comprised 200 valid blood smear samples, corresponding to 200 distinct patients, with no missing values in the analyzed variables. Four erythrocyte parameters were evaluated: RBC, HGB, HCT, and RDW-CV. Descriptive statistics for these parameters are presented in Table 2.

**Table 2:** Erythrocyte hematologic parameters of the analyzed samples.

| Parameter | Mean ± SD | Median; IQR | Range | Unit |
| --- | --- | --- | --- | --- |
| RBC | $4.34 \pm 0.74$ | 4.40; 3.90–4.81 | 2.10–6.04 | $10^6/\mu\text{L}$ |
| HGB | $12.94 \pm 2.14$ | 13.10; 11.98–14.30 | 6.8–17.5 | g/dL |
| HCT | $38.13 \pm 5.98$ | 38.75; 35.33–41.93 | 20.1–51.3 | % |
| RDW-CV | $13.36 \pm 2.09$ | 12.80; 12.10–13.73 | 10.9–24.8 | % |

#### 2.6.2 Data Distribution

Figure 2 presents the histograms, indicating that RBC, HGB, and HCT exhibit approximately normal distributions, with close means and medians. In contrast, RDW-CV showed positive skewness, with a concentration of values below the mean and a longer right tail.

**Figure 2:**
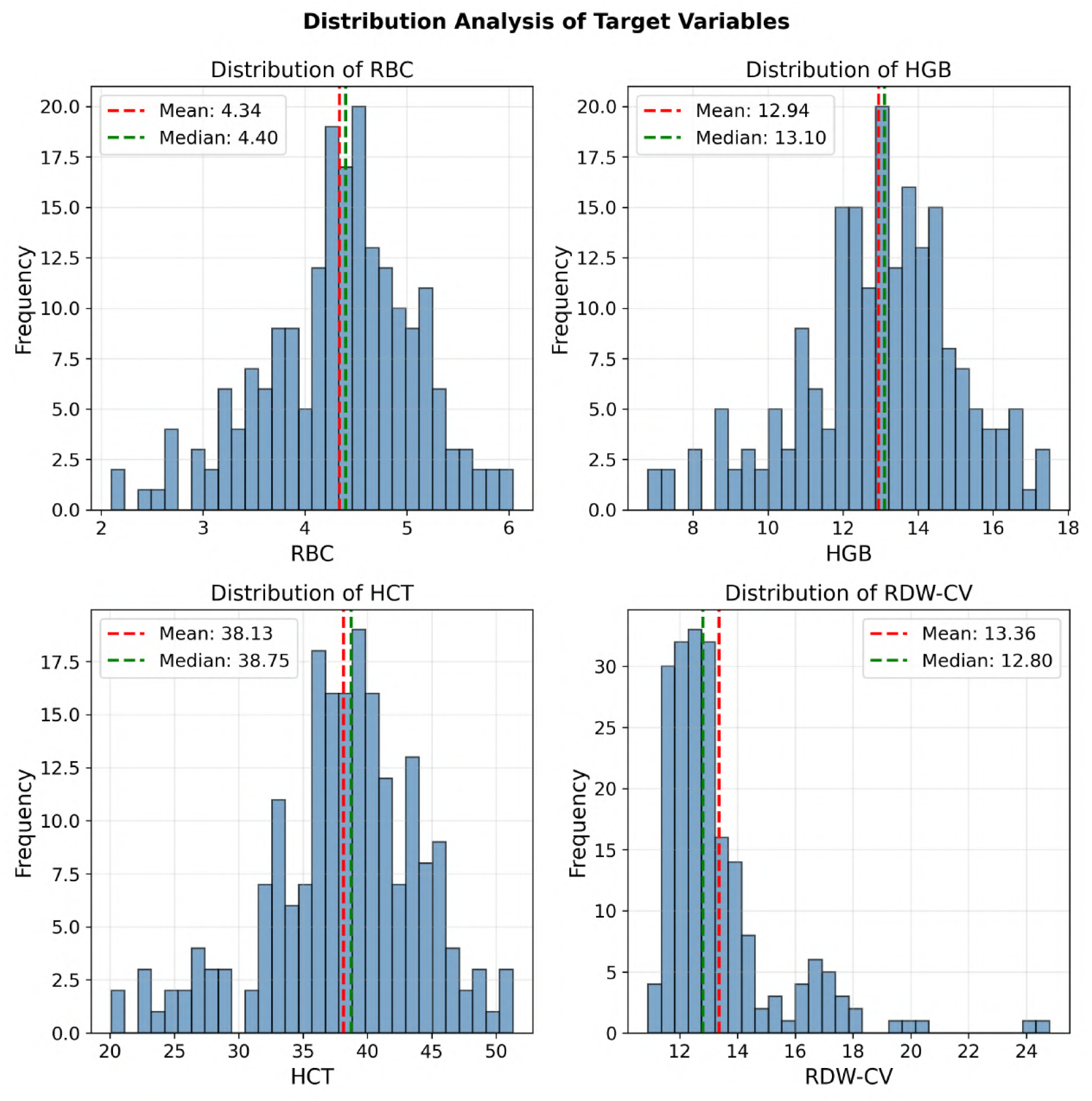
Distribution of erythrocyte hematological parameters. Histograms showing the observation frequency for RBC, HGB, HCT, and RDW-CV, with indication of mean in red dashed line and median in green dashed line.

Figure 3 presents the boxplots of the analyzed parameters, indicating the presence of outliers in all variables. Three outliers were identified in RBC, 7 in HGB, 8 in HCT, and 24 in RDW-CV. Outlier detection was performed using the interquartile range method, IQR, as described by Tukey [7].

**Figure 3:**
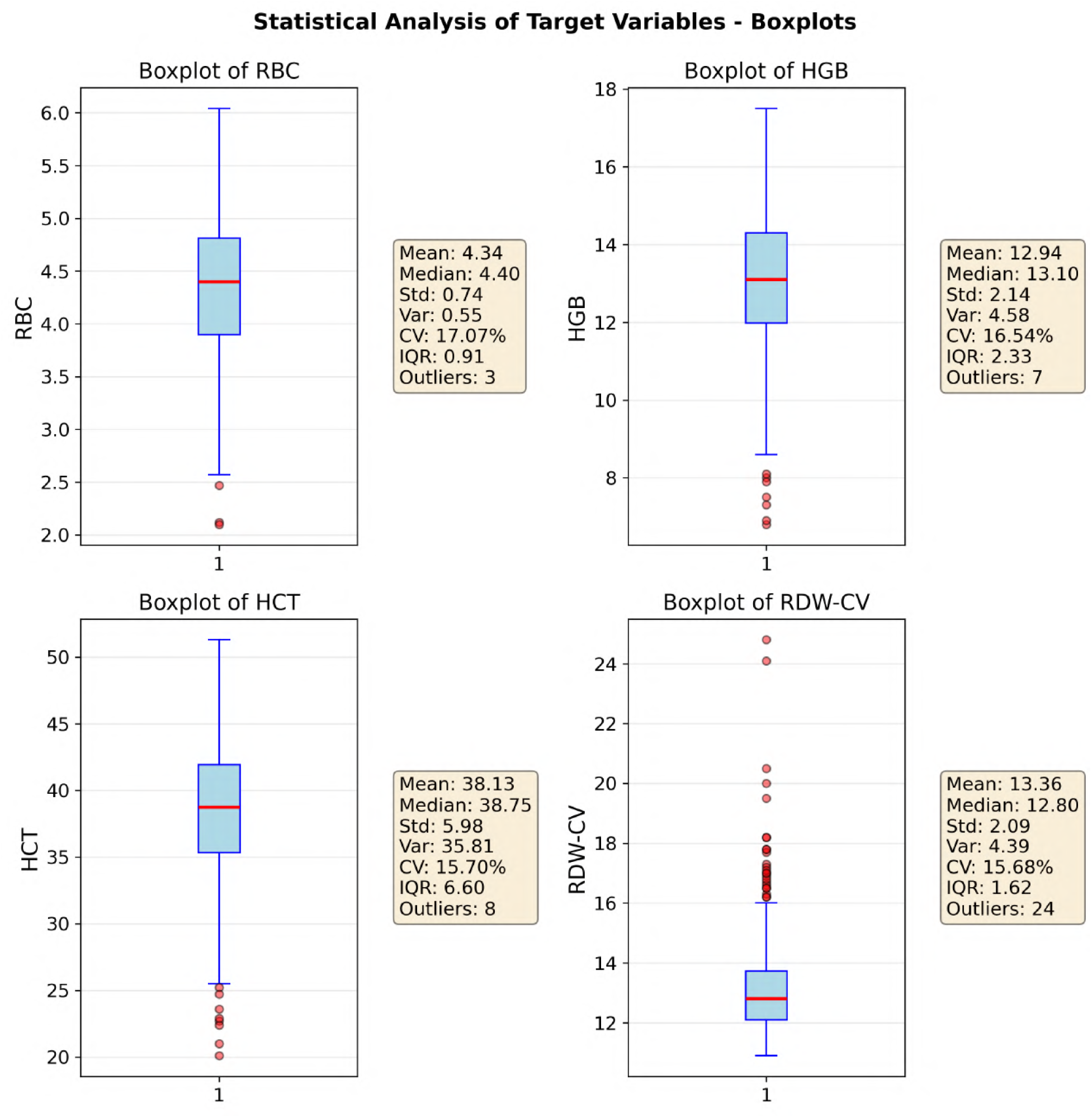
Boxplots of erythrocyte hematological parameters. Graphical representation showing median, interquartile ranges and outliers highlighted in red dots for RBC, HGB, HCT, and RDW-CV. To the right of each boxplot, a panel with the metrics for that parameter.

#### 2.6.3 Correlation Analysis

Figure 4 presents the Pearson correlation matrix, *r*, among the erythrocyte parameters, highlighting the patterns of linear association among hematologic variables [8]. A strong positive correlation was observed among RBC, HGB, and HCT, with *r* = 0.919 between RBC and HGB, *r* = 0.926 between RBC and HCT, and *r* = 0.982 between HGB and HCT. In contrast, RDW-CV showed moderate negative correlations with the other parameters, ranging from *r* = *−*0.534 with RBC to *r* = *−*0.635 with HGB.

**Figure 4:**
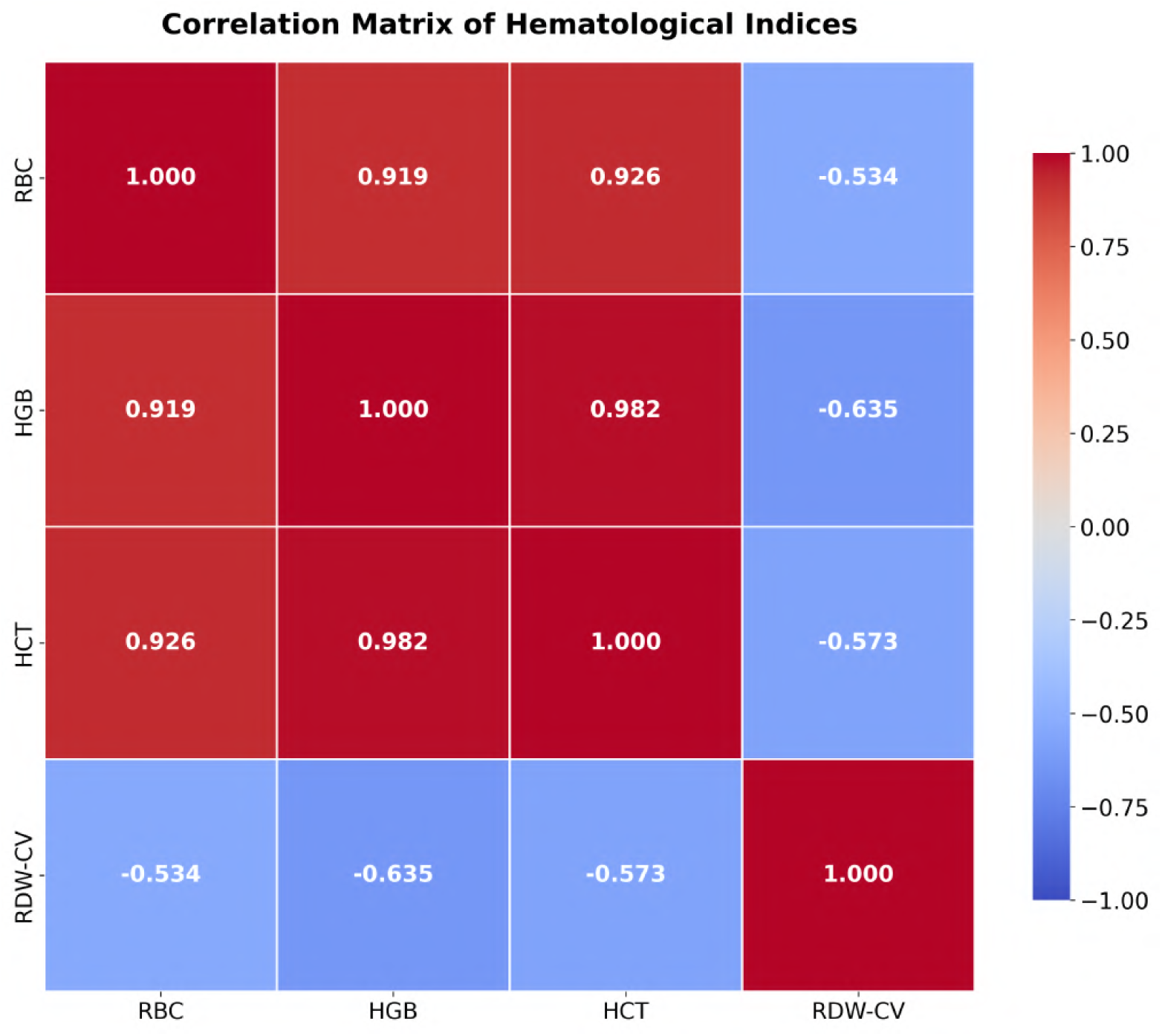
Correlation matrix between erythrocyte hematological parameters. Heat map representing Pearson correlations, where red indicates strong positive correlation and blue indicates negative correlation.

#### 2.6.4 Principal Component Analysis, PCA

To identify variability patterns among hematologic parameters, PCA was applied [9]. The variance decomposition shown in Figure 5 indicates that the first two principal components account for 97% of the total variance, with 83% attributed to the first component and 14% to the second.

**Figure 5:**
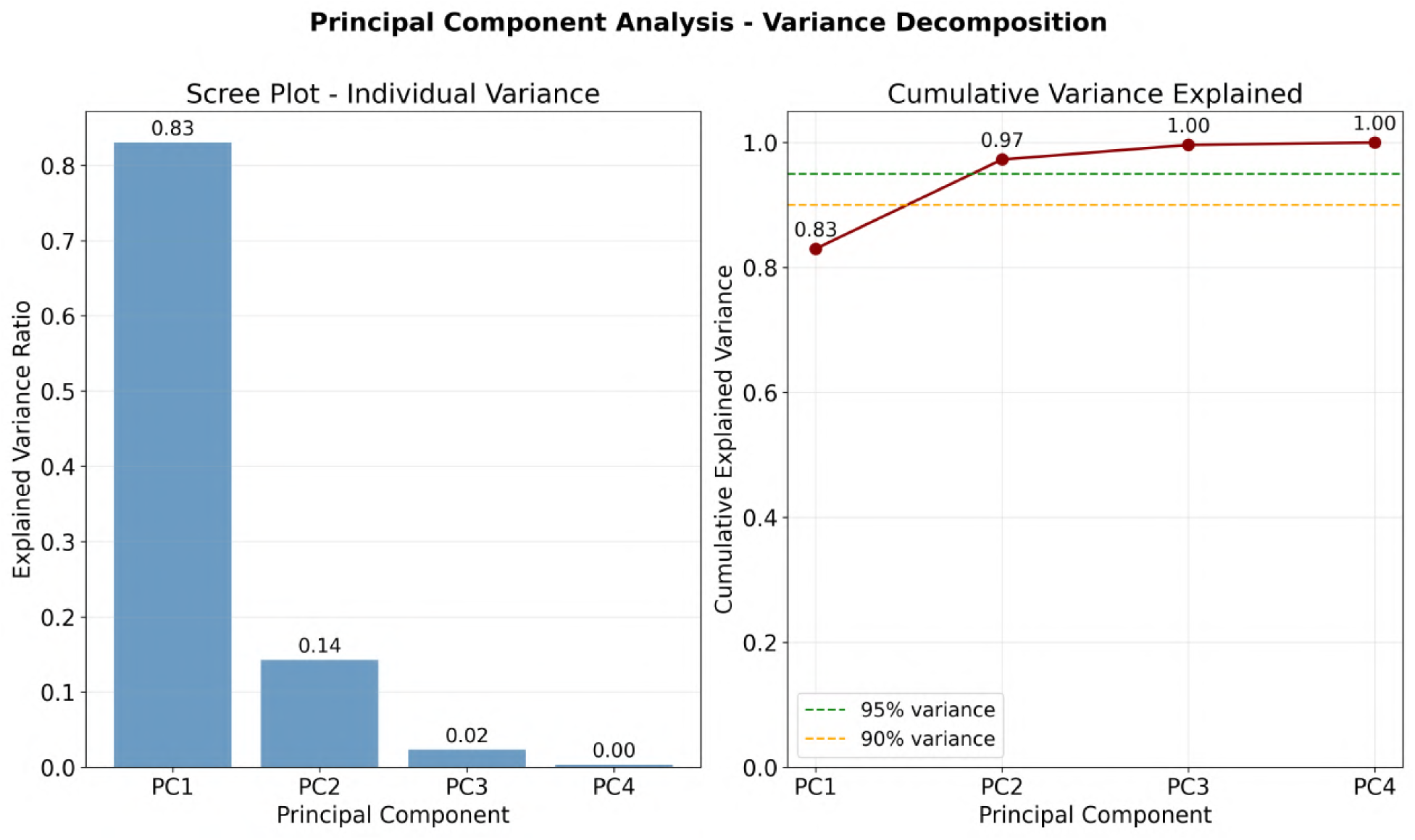
Variance decomposition explained by PCA. Left, individual variance of each principal component. Right, cumulative explained variance.

The PCA biplot in Figure 6 illustrates the distribution of samples and variable contributions in the first two components. RBC, HGB, and HCT show vectors with similar direction and magnitude, loading strongly on PC1. In contrast, RDW-CV exhibits a different direction, with higher loading on PC2, reflecting its different relationship with the other parameters.

**Figure 6:**
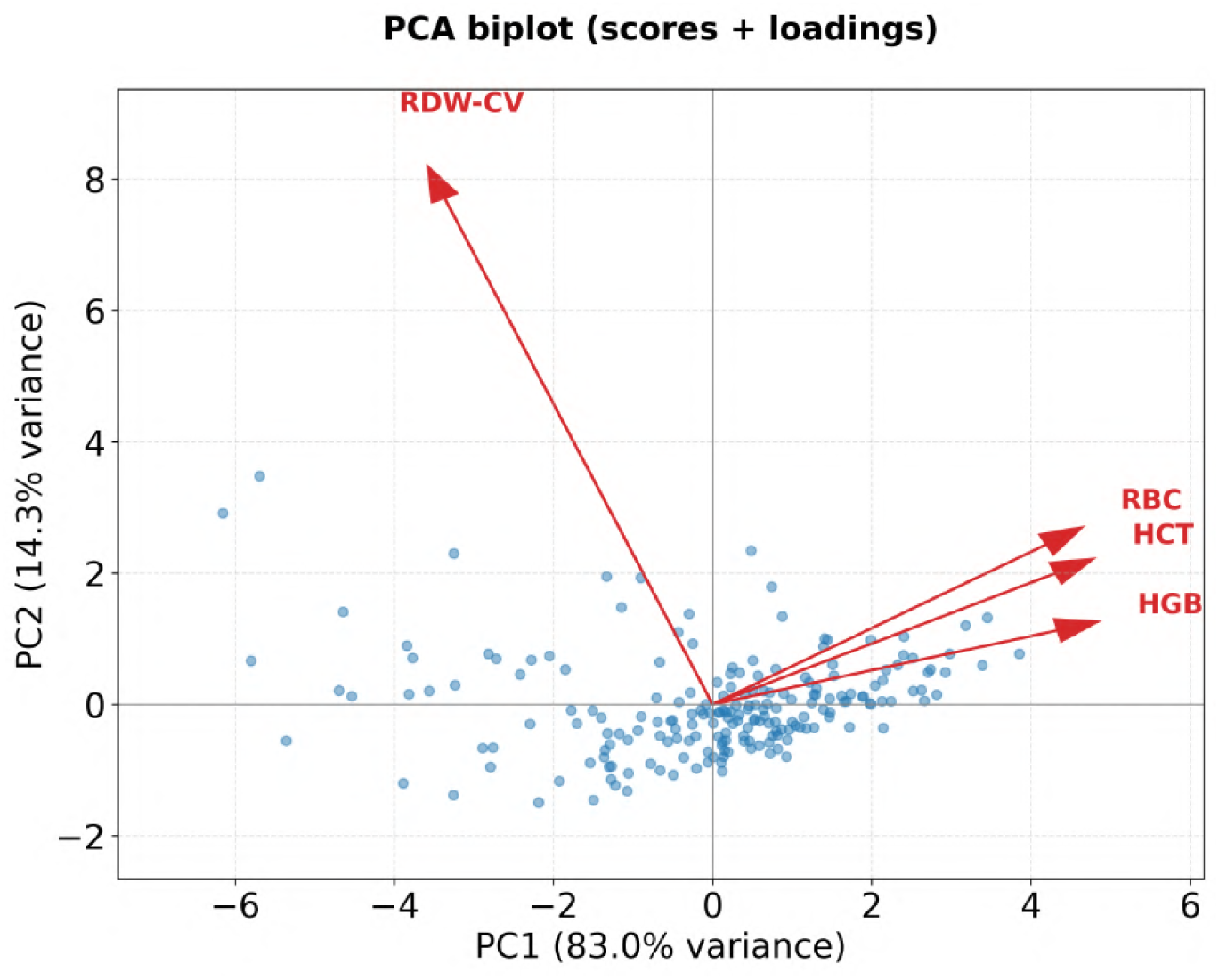
Biplot of principal component analysis. Distribution of samples in blue dots and contribution of variables in red arrows in the first two principal components.

#### 2.6.5 Experimental Design

Based on the PCA biplot in Figure 6, two main axes of variability were identified. RBC, HGB, and HCT showed nearly colinear loadings on PC1, indicating strong interdependence. This justified the use of a multi-output model that leverages this relationship among variables. RDW-CV loaded mainly on PC2 and showed independent behavior, motivating the use of a dedicated single-output model. This PCA-guided division makes the process more efficient and reduces the number of required experiments by half, without loss of statistical consistency.

Four convolutional neural network architectures were selected because they combine established relevance in clinical applications and computational feasibility: ResNet-18 and ResNet-34, based on residual connections with established performance in hematologic analysis [10]; DenseNet-121, effective on limited datasets due to intensive feature reuse [11]; and EfficientNet-B0, with a good performance–cost trade-off via compound scaling [12]. The choice also considered the limitations of the available hardware, thereby rendering larger models impractical. All architectures were initialized with ImageNet pretrained weights (IMAGENET1K_V1), leveraging transfer learning from a large natural image dataset [13], which promotes faster and improved initial generalization in the blood smear domain.

Each architecture was evaluated at four image resolutions, 224, 512, 768, and 1024 pixels, to assess the impact of resolution on predictive performance, resulting in a total of 32 distinct experiments, considering two model groups, four image resolutions, and four architectures. Pre-trained networks were adapted for regression tasks by replacing their final classification layers with a custom regression block, in which num_features corresponds to the number of features extracted by the backbone and num_outputs depends on the prediction group, with 3 outputs for RBC, HGB, and HCT and 1 output for RDW-CV, as shown in Figures 7 and 8.

**Figure 7:**
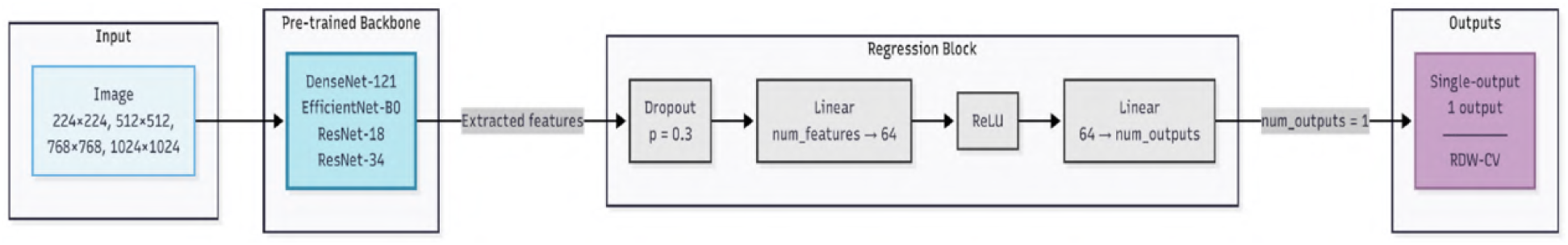
Proposed model architecture, illustrating the processing flow from image input to the regression block and the single-output prediction.

**Figure 8:**
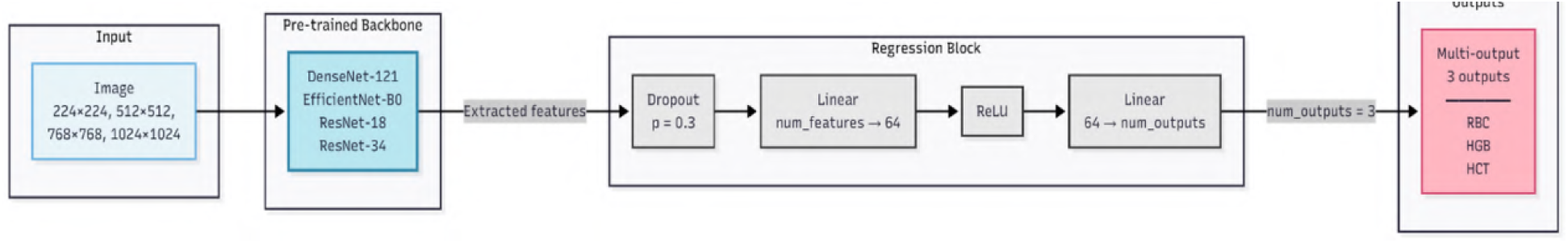
Proposed model architecture, illustrating the processing flow from image input to the regression block and the multi-output prediction.

Given the limited size of the dataset, K-Fold cross-validation with *k* = 10 and random_state = 100 was used to evaluate the models. This strategy avoids dependence on a single random split between training and validation and provides more robust and generalizable estimates of performance metrics [14].

In this process, numerical data were standardized using StandardScaler, which scales each variable to zero mean and unit variance. This standardization reduces scale differences among variables and contributes to more stable and efficient training, improving optimizer behavior and facilitating faster convergence [15]. The procedure was correctly applied in each fold, using fit_transform only on the training set and transform on the validation set, avoiding data leakage [16].

In addition, the training image set was augmented through vertical and horizontal flipping and 180° rotation, quadrupling the number of available samples from 180 to 720 images. This data augmentation step is essential due to the small size of the original dataset, helping mitigate overfitting and improving model generalization [17]. Data augmentation was applied exclusively to the training images after partitioning in each fold to avoid data leakage [16].

Models were optimized using hyperparameters tuned for fine-tuning on small datasets. The learning rate was chosen to allow the adaptation of pre-trained backbones without abruptly overwriting the representations learned on ImageNet [18]. Weight decay follows classic recommendations to improve generalization in deep networks [19]. The adopted loss function was Huber Loss (*δ* = 1), which is less sensitive to outliers than MSE and more stable than MAE during gradient-based optimization [20], an important characteristic given the outliers identified in the exploratory analysis.

Dropout was used in the regression head to reduce neuron coadaptation, and the Adam optimizer was used due to its stability at low learning rates. The number of epochs was defined because pretrained networks tend to converge quickly in low-sample settings, requiring only fine-tuning of the weights to adapt to the new domain. Batch size varied across experiments according to image resolution, respecting GPU memory limits and maintaining VRAM utilization close to 83% to maximize hardware usage [21].

**Table 3:** Hyperparameters used in model training.

| Hyperparameter | Value |
| --- | --- |
| Learning rate | $5 \times 10^{-5}$ |
| Weight decay | $1 \times 10^{-4}$ |
| Loss function | Huber Loss ($\delta = 1$) |
| Dropout | 0.3 |
| Optimizer | Adam |
| Epochs | 50 |
| Batch size | Adjusted |

All experiments were conducted locally in an environment equipped with an NVIDIA GeForce RTX 4070 Ti GPU with 12GB VRAM, an AMD Ryzen7 5800X3D processor with 8 cores, 3.4GHz, and 32GB RAM.

## 3 Results

### 3.1 Selection of the Best Models

Among the experiments evaluated, those with the lowest mean absolute error, MAE, were selected, a metric chosen for its robustness to extreme values and its direct interpretability on the clinical scale [22], particularly relevant given the outliers identified in the exploratory analysis, as shown in Figure 3. As a complementary measure, the coefficient of determination, *R*^2^, was used, which quantifies the proportion of variability explained by the predictions [23].

In addition to MAE and *R*^2^ means, 95% confidence intervals, 95% CI, were computed from the ten cross-validation folds. Inclusion of 95% CIs allows quantification of variability across partitions and the uncertainty associated with performance, as recommended in statistical evaluations of models based on cross-validation [24].

As illustrated in Figure 9, which presents performance curves for different models as a function of image size for RBC, HGB, and HCT, the best performance was achieved by ResNet-34 using 1024*×*1024 pixel images. The mean values for this configuration, *MAE* = 0.667 and *R*^2^ = 0.892, computed from cross-validation, are presented in Table 4. These results were followed by ResNet-18 at the same resolution, with *MAE* = 0.693 and *R*^2^ = 0.889.

**Figure 9:**
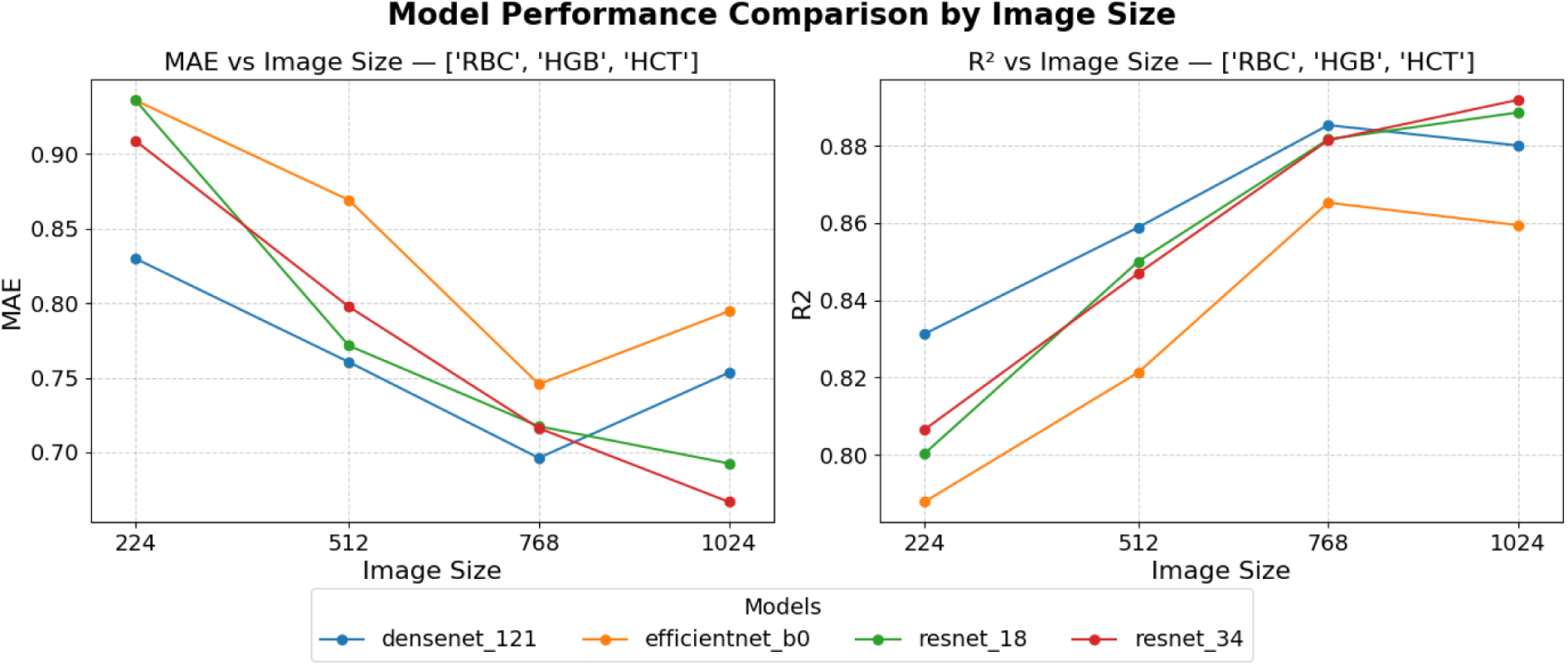
Performance of convolutional models as a function of image size for RBC, HGB, and HCT. Left, MAE; right, *R*^2^.

**Table 4:** Cross-validation metrics for the ResNet-34 model with 1024*×*1024 images.

| Result | MAE | SD | CI 95% | $R^2$ | SD | CI 95% |
| --- | --- | --- | --- | --- | --- | --- |
| **Overall (between folds)** | **0.667** | **0.105** | **±0.075** | **0.892** | **0.061** | **±0.044** |
| RBC | 0.184 | 0.035 | ±0.025 | 0.889 | 0.042 | ±0.030 |
| HGB | 0.524 | 0.092 | ±0.066 | 0.874 | 0.108 | ±0.077 |
| HCT | 1.292 | 0.229 | ±0.164 | 0.912 | 0.051 | ±0.037 |

For RDW-CV, the best result based on MAE was obtained by DenseNet-121 using 1024*×*1024 pixel images, achieving *MAE* = 0.960 and *R*^2^ = 0.435. This was followed by EfficientNet-B0 using 768*×*768 pixel images, achieving *MAE* = 0.966 and *R*^2^ = 0.491, as shown in Figure 10. The corresponding mean metrics for these configurations are presented in Table 5.

**Figure 10:**
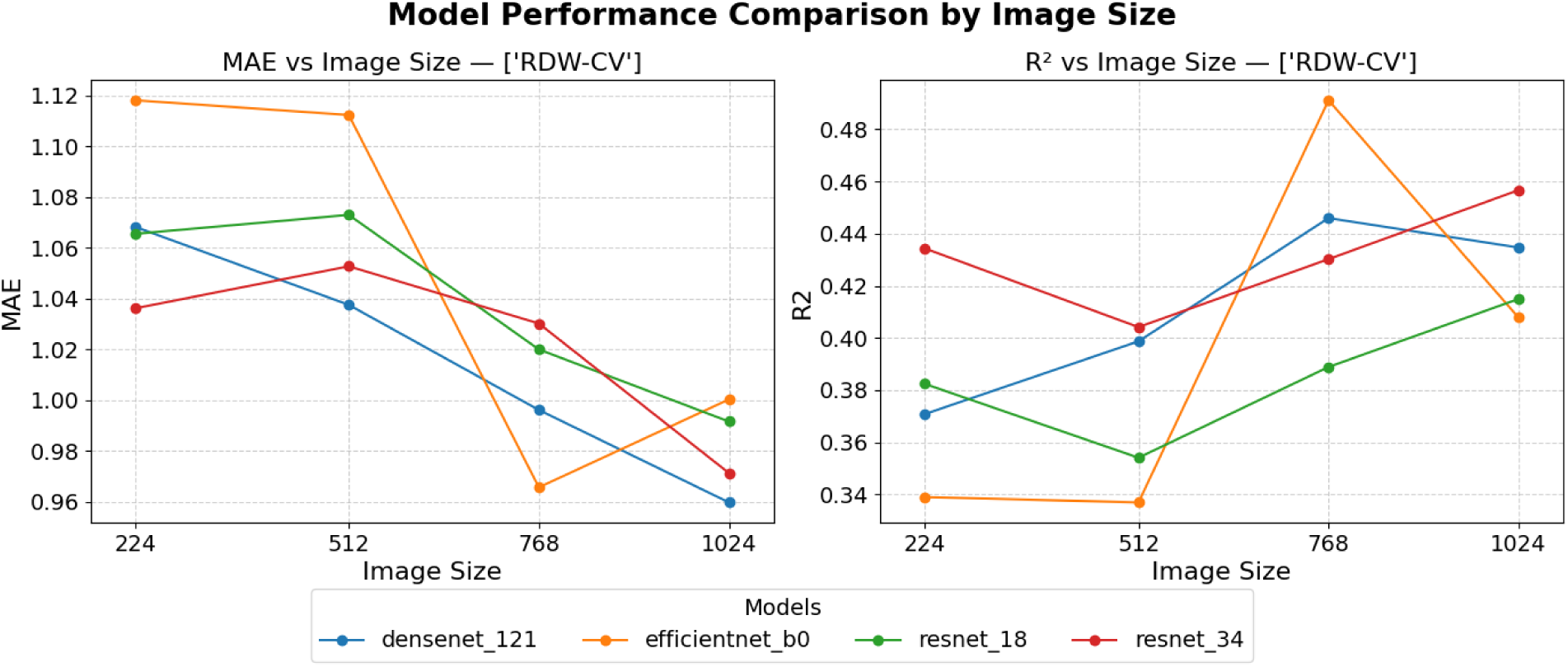
Performance of convolutional models as a function of image size for RDW-CV. Left, MAE; right, *R*^2^.

**Table 5:** Cross-validation metrics for the DenseNet-121 model with 1024*×*1024 images.

| Result | MAE | SD | CI 95% | $R^2$ | SD | CI 95% |
| --- | --- | --- | --- | --- | --- | --- |
| Overall (between folds) | 0.960 | 0.323 | $\pm 0.231$ | 0.435 | 0.230 | $\pm 0.165$ |

### 3.2 Training Curves

Training was performed for 50 epochs across each of the 10 cross-validation folds, totaling 500 epochs. The ResNet-34 architecture for RBC, HGB, and HCT required 267.1 minutes, with a mean time per epoch of 32.06 s, whereas DenseNet-121 for RDW-CV required 452.3 minutes, with a mean of 54.28 s per epoch. Training curves shown in Figures 11 and 12 indicate stable convergence, with the best-performing models across folds identified around epochs 24.5 *±* 15.3 for ResNet-34 and 23.9 *±* 15.5 for DenseNet-121, varying between epochs 5–47 and 4–46, respectively. The val_loss/train_loss ratios of 1.69 and 2.00 confirm that both models are adequately regularized, with no significant signs of overfitting, highlighting the effectiveness of the applied regularization techniques.

**Figure 11:**
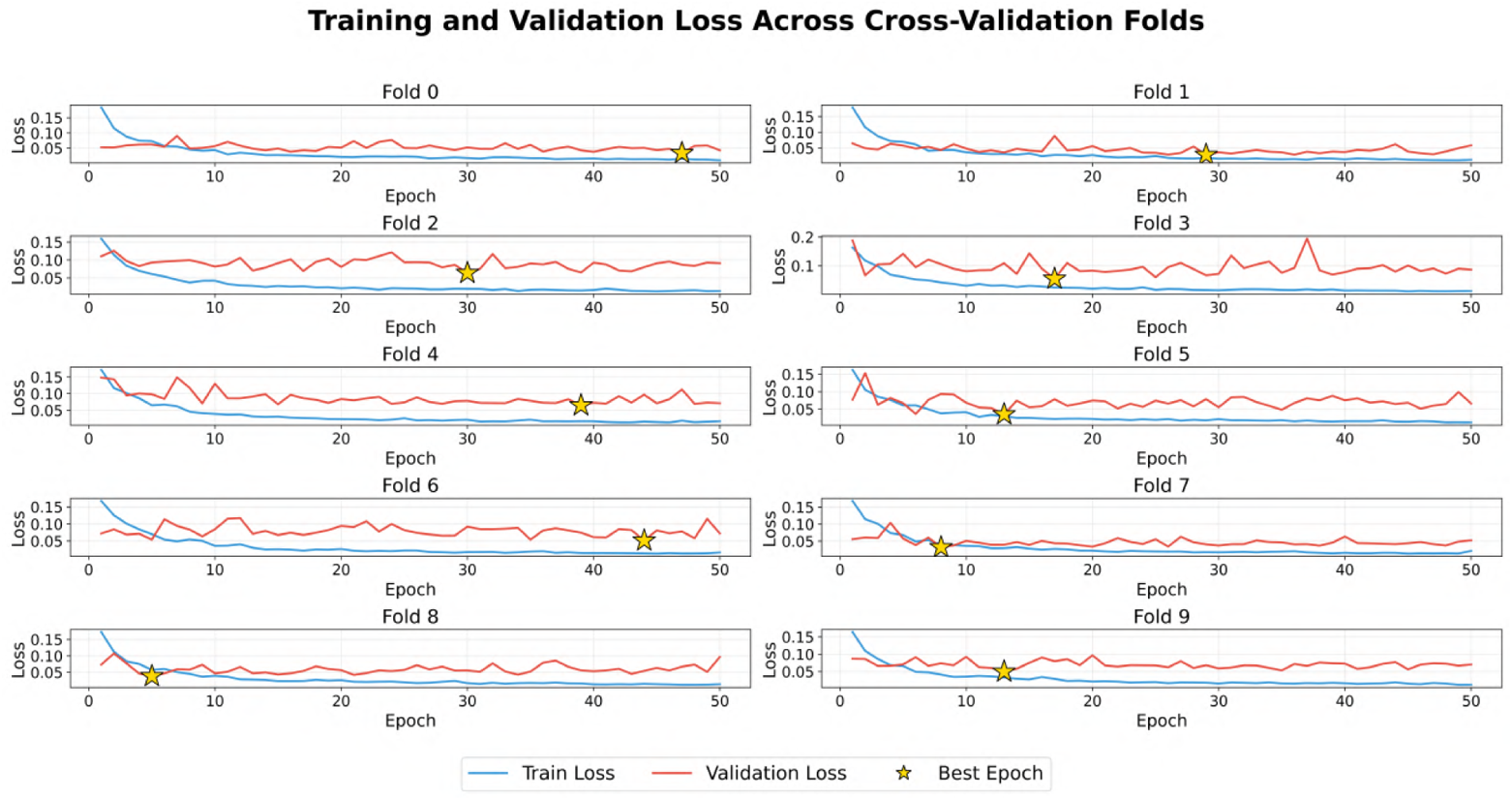
Training and validation curves per fold for ResNet-34 model. Evolution of loss function over epochs for each of the 10 cross-validation folds. Yellow stars indicate the epoch of best performance.

**Figure 12:**
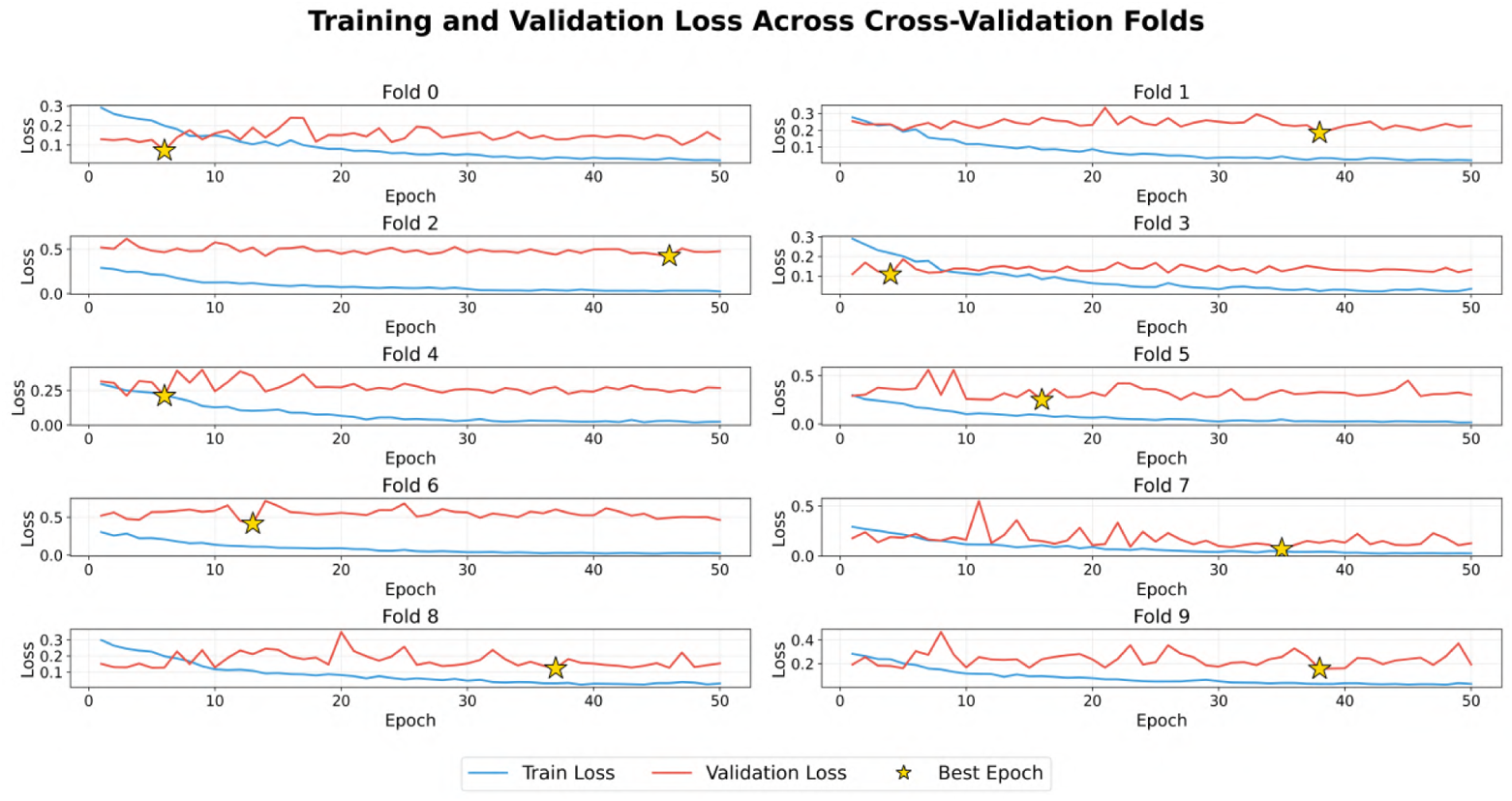
Training and validation curves per fold for DenseNet-121 model. Evolution of loss function over epochs for each of the 10 cross-validation folds. Yellow stars indicate the epoch of best performance.

### 3.3 Performance by Analyte

To evaluate analyte-specific performance for the selected model, samples from the corresponding validation fold were used, ensuring that no sample was evaluated by a model that had used it during training. Reported metrics, *R*^2^, MAE, root mean squared error, RMSE, and Pearson correlation, *r*, were computed by aggregating all predictions obtained on the folds’ validation sets, providing a robust estimate of model performance for each analyte. RMSE was included due to its higher sensitivity to large deviations. By penalizing large errors that MAE may not reveal, RMSE provides a more conservative estimate of model performance [22].

Figure 13 presents scatter plots of true versus predicted values for each analyte, allowing visualization of the relationship between model predictions and laboratory reference values. Table 6 summarizes the performance metrics obtained for primary analytes and derived indices calculated from model predictions.

**Figure 13:**
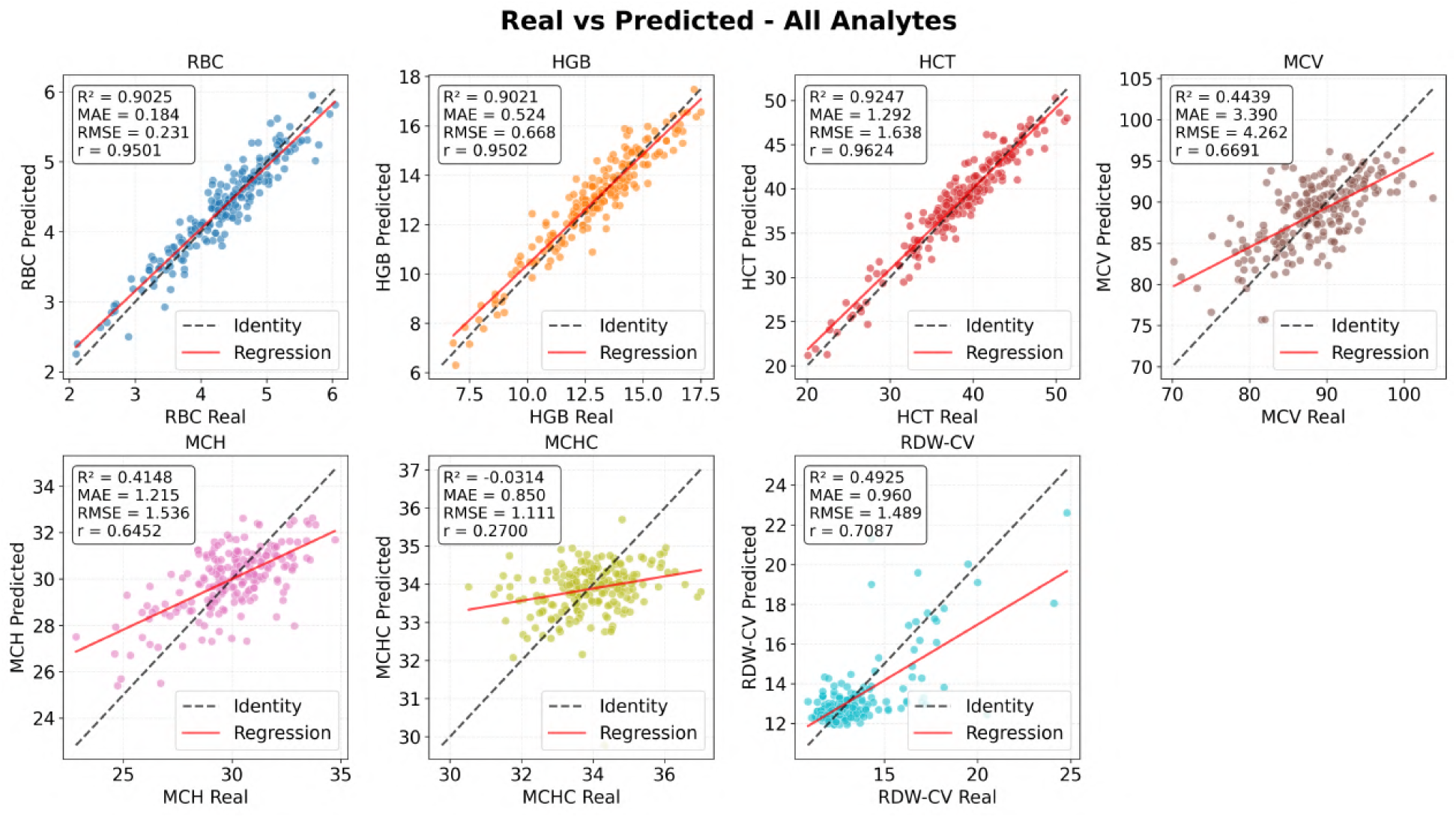
Relationship between real and predicted values for all analytes. Scatter plots showing the correlation between reference values on x-axis and model-predicted values on y-axis. Black dashed line represents perfect identity; red line represents the fitted linear regression.

**Table 6:**
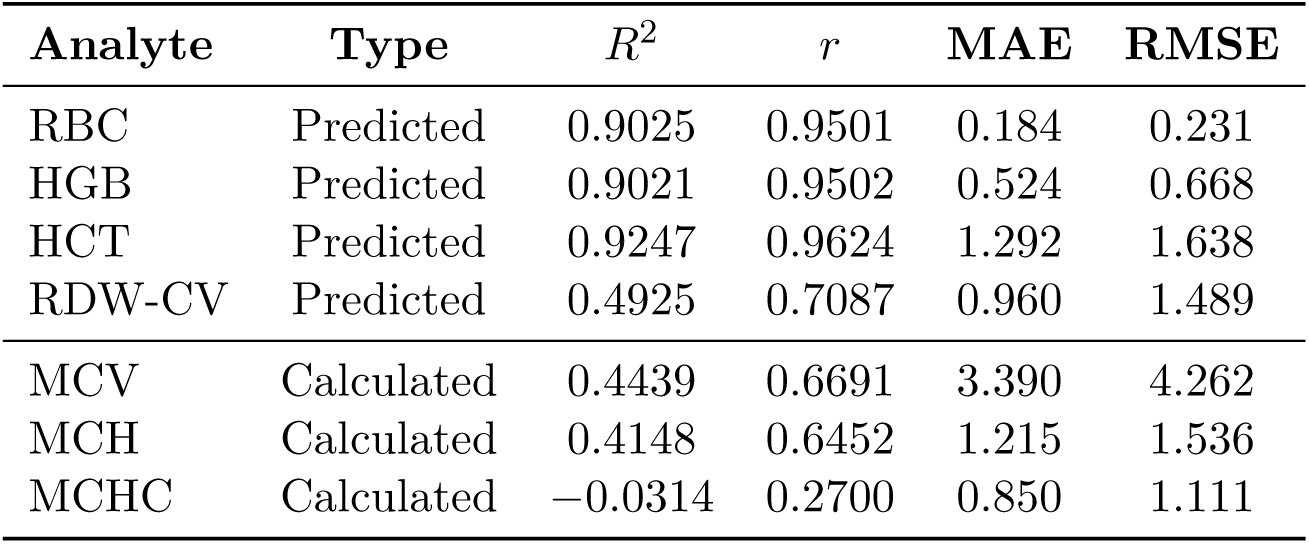
Performance metrics for primary analytes and derived indices calculated from model predictions.

| Analyte | Type | $R^2$ | $r$ | MAE | RMSE |
| --- | --- | --- | --- | --- | --- |
| RBC | Predicted | 0.9025 | 0.9501 | 0.184 | 0.231 |
| HGB | Predicted | 0.9021 | 0.9502 | 0.524 | 0.668 |
| HCT | Predicted | 0.9247 | 0.9624 | 1.292 | 1.638 |
| RDW-CV | Predicted | 0.4925 | 0.7087 | 0.960 | 1.489 |
| MCV | Calculated | 0.4439 | 0.6691 | 3.390 | 4.262 |
| MCH | Calculated | 0.4148 | 0.6452 | 1.215 | 1.536 |
| MCHC | Calculated | -0.0314 | 0.2700 | 0.850 | 1.111 |

The predicted values for RBC, HGB, and HCT showed excellent performance, with *R*^2^ values between 0.90 and 0.92, *r* above 0.95, and MAE values of 0.184, 0.524, and 1.292, respectively. RDW-CV exhibited intermediate performance, with *R*^2^ = 0.49, *r* = 0.71, and MAE = 0.96. The derived indices, MCV and MCH showed lower *R*^2^ values of 0.44 and 0.41, respectively, while maintaining *r* = 0.67 and 0.65. In contrast, MCHC exhibited inferior performance, with *R*^2^ values close to zero and *r* = 0.27.

### 3.4 Grad-CAM Visualization and Interpretation

Grad-CAM visualizations [25], Figures 14 and 15, show that for RBC, HGB, and HCT, activations are mainly concentrated in regions with higher erythrocyte density. For RDW-CV, the maps exhibit more diffuse and less spatially defined activations. These patterns highlight differences in prediction stability among analytes and in the consistency of regions considered relevant by the model.

**Figure 14:**
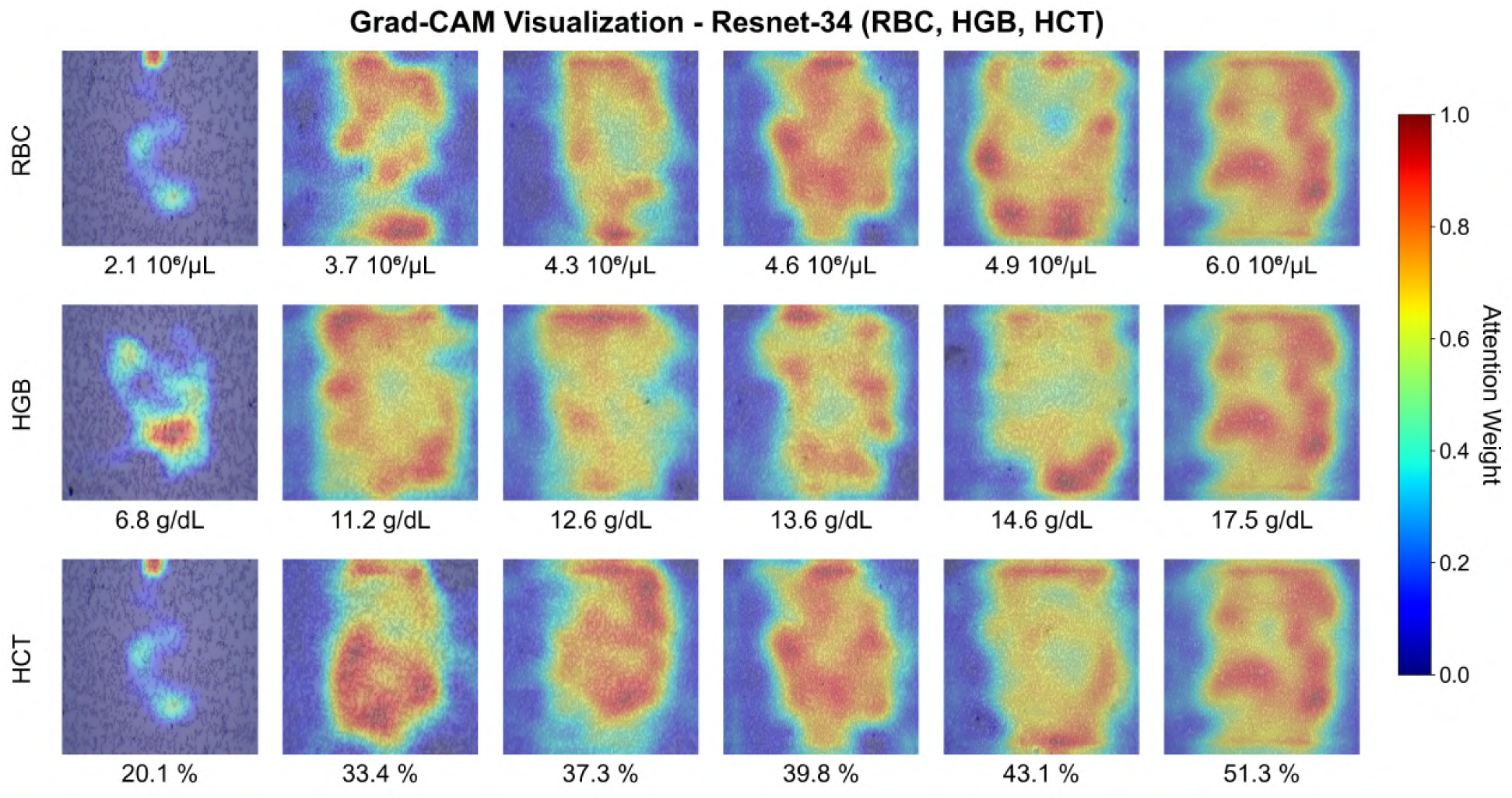
Grad-CAM visualization for RBC, HGB and HCT. Attention maps of ResNet-34 model overlaid on original images, where warm colors indicate regions of greater importance for prediction.

**Figure 15:**
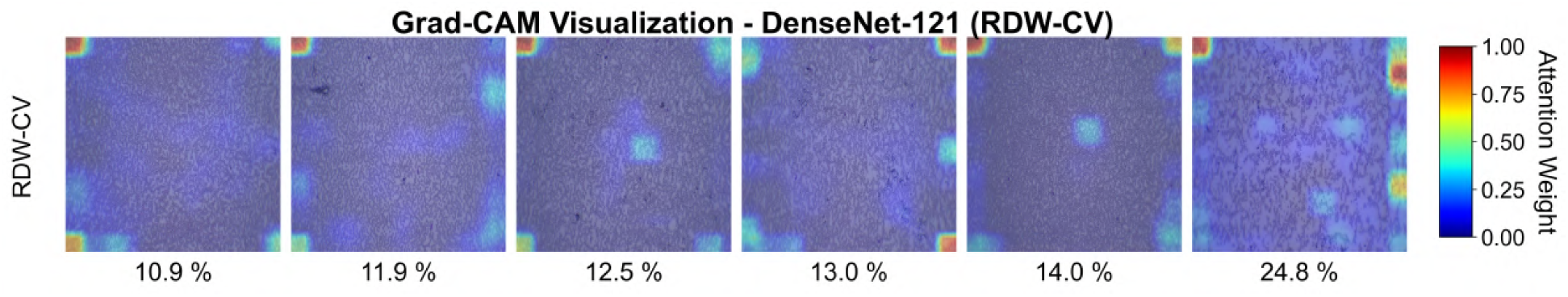
Grad-CAM visualization for RDW-CV. Attention maps of DenseNet-121 model overlaid on original images, where warm colors indicate regions of greater importance for prediction.

### 3.5 Bland–Altman Agreement Analysis

Bland–Altman analysis, shown in Figure 16, demonstrated mean biases close to zero for all predicted parameters. For RBC, HGB, HCT, and RDW-CV, biases were 0.006 *×* 10^6^*/µ*L, 0.059 g/dL, 0.21%, and *−*0.11%, respectively. The 95% limits of agreement ranged from *−*0.447 to 0.459 *×* 10^6^*/µ*L for RBC, *−*1.249 to 1.367 g/dL for HGB, *−*2.979 to 3.403% for HCT, and *−*3.028 to 2.805% for RDW-CV. Between 94% and 95% of observations remained within these limits. For derived indices, biases were: MCV = 0.198 fL, MCH = 0.035 pg, and MCHC = *−*0.039 g/dL, with limits of agreement of *−*8.169 to 8.564 fL, *−*2.982 to 3.053 pg, and *−*2.221 to 2.143 g/dL. The proportion of observations within the limits ranged from 94.5% to 95.5%. No parameter showed a systematic pattern of variation across the measurement range, indicating absence of proportional bias.

**Figure 16:**
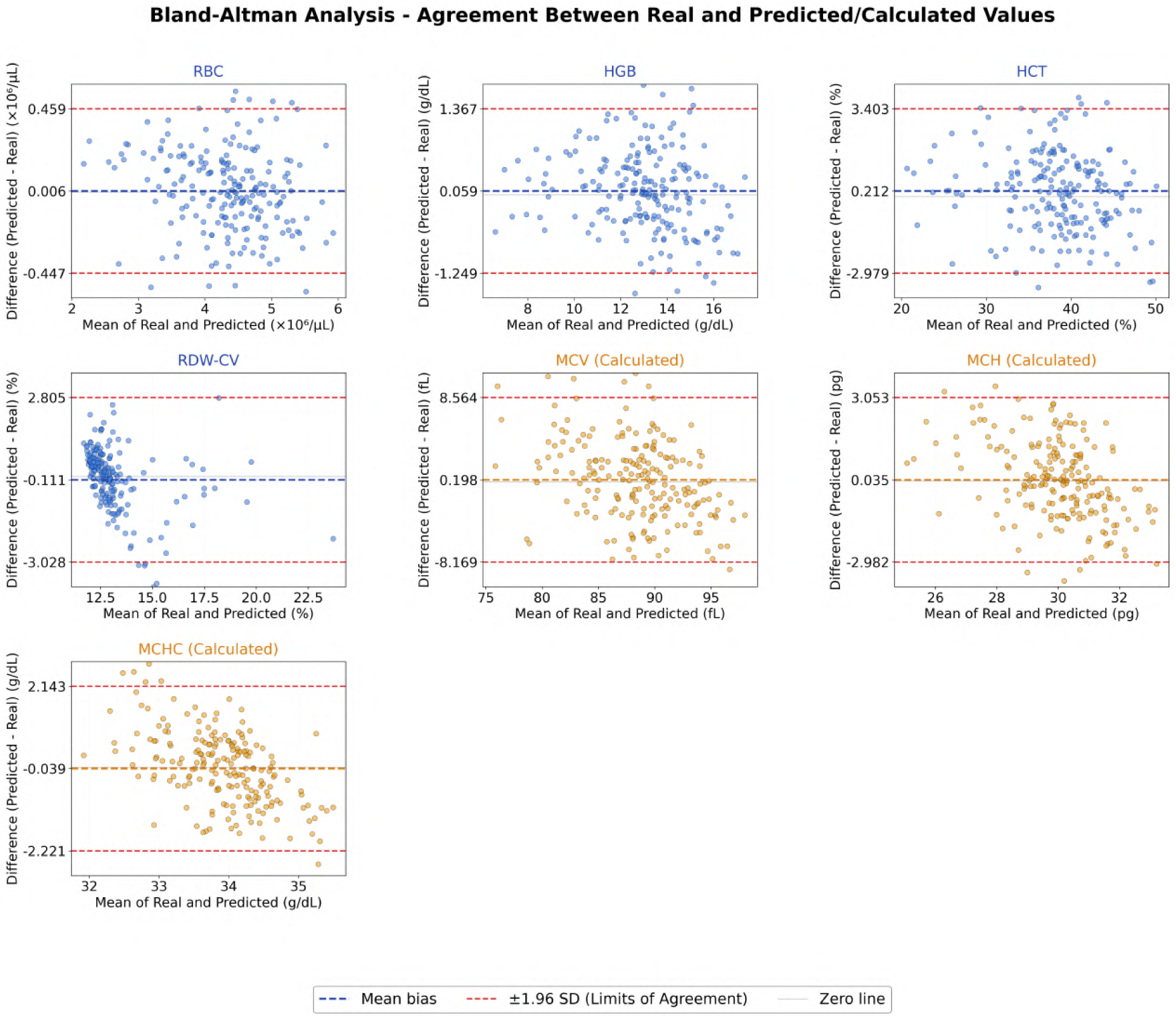
Bland–Altman analysis of erythrocyte parameters. Graphs showing the difference between predicted and real values as a function of the mean of the two methods. Orange dashed line represents mean bias; red dashed lines represent 95% limits of agreement; gray line represents zero difference.

## 4 Discussion

The erythrogram is fundamental for hematologic assessment, but its acquisition remains dependent on automated analyzers; to the best of current knowledge, no studies have investigated direct estimation of its indices from images obtained through the eyepiece of conventional optical microscopes, despite recent advances in artificial intelligence applied to blood smears.

In this context, the models evaluated in this study showed excellent performance in predicting the primary analytes, RBC, HGB, and HCT, with *R*^2^ above 0.90, *r* above 0.95, and low MAE and RMSE, indicating agreement between true and predicted values. HCT showed the best performance with *R*^2^ = 0.92 and *r* = 0.96, followed by HGB and RBC with *R*^2^ *≈* 0.90 and *r ≈* 0.95, indicating high predictive accuracy and consistent identification of morphological patterns associated with variations in the red cell series.

RDW-CV showed intermediate performance with *R*^2^ = 0.49 and *r* = 0.71, as expected given its greater sensitivity to subtle morphological variations. Despite the higher observed variability, the model maintained a consistent linear trend between true and predicted values, indicating that part of the variability was correctly represented.

In contrast, derived indices, MCV, MCH, and MCHC, showed lower performance with *R*^2^ between *−*0.03 and 0.44, as expected due to their direct dependence on predictions of the primary analytes. These parameters were not predicted by the model, but calculated from RBC, HGB, and HCT outputs according to Equations 1, 2, and 3:

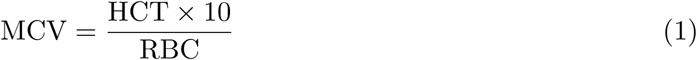

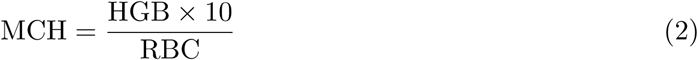

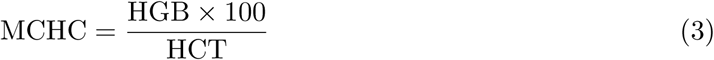

These formulas involve ratio operations between correlated variables, which amplifies the propagation of errors from the input estimates. Small inaccuracies in HCT or RBC, for example, result in disproportionately larger variations in MCV and MCH. This cumulative effect explains the reduction in *R*^2^ and the increase in the observed mean squared error in the derived indices.

MCHC showed an *R*^2^ value close to zero, attributed to its narrow dynamic range, approximately between 30 and 37 g/dL, which limits the total explainable variance. In such cases, even small absolute deviations can substantially reduce statistical explanatory power without necessarily compromising the clinical relevance of the predictions.

### 4.1 Analytical Agreement

Bland–Altman analysis demonstrated good analytical agreement between predicted and reference values for the primary parameters, including RBC, HGB, HCT, and RDW-CV, with biases close to zero and more than 94.0% of observations within the limits of agreement. The absence of proportional bias confirms the consistency of the estimates across the measurement range. The greater dispersion observed for RDW-CV reflects its sensitivity to subtle morphological variations. Derived indices, MCV, MCH, and MCHC, also showed biases close to zero and an adequate proportion of observations within the limits of agreement between 94.5% and 95.5%. However, the proportionally wider limits observed for these indices reflect cumulative propagation of errors from primary predictions, as expected for ratio operations between correlated variables. This finding reinforces that direct prediction of these indices, rather than calculation from primary outputs, could improve performance, as suggested in the limitations.

Identified outliers, although infrequent, may be related to the small sample size, suggesting that expanding the dataset may reduce their influence and improve the robustness of the results.

### 4.2 Comparison with the study by Yang et al., 2025

Values reported by Yang et al. [6] were originally presented in the International System of Units, g/L, L/L, and 10^12^/L. To enable direct comparison with the data from this study, these values were converted to the conventional units used here according to Equations 4, 5, and 6:

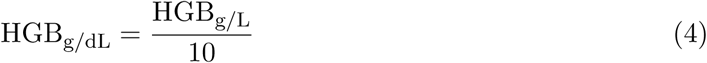

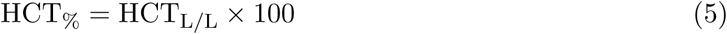

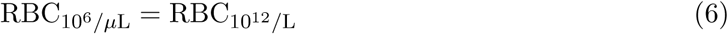

Despite differences in scale and resources, a strong convergence in the results is observed. This work used 200 samples from a single laboratory and achieved *R*^2^ between 0.90 and 0.92 and *r >* 0.95 for RBC, HGB, and HCT, which are close to those reported by Yang et al. [6], who used 13,592 images from three institutions and automated laboratory infrastructure such as the CellaVision DI-60 and Sysmex XN-9000 systems and obtained *R*^2^ values between 0.95 and 0.96 for the same parameters.

Models developed in this study showed comparable performance, as evidenced by the MAE values obtained for the primary analytes: *RBC* = 0.18, *HGB* = 0.52, and *HCT* = 1.29, when compared with those reported by Yang et al. [6]: *RBC* = 0.16, *HGB* = 0.46, and *HCT* = 1.30. These results suggest that high accuracy can be achieved using accessible resources, widely available equipment, and multi-output models. This expands the potential application of the method in laboratory settings with lower infrastructure, which can contribute to broader democratization of access to healthcare.

Regarding RDW-CV, both studies showed lower performance compared to primary analytes. In this work, the model achieved *R*^2^ = 0.49, while Yang et al. [6] reported *R*^2^ = 0.88, a difference attributed to the greater sensitivity of RDW-CV to subtle morphological variations and to the smaller number of samples available for training. Even so, the model maintained a stable linear trend and good generalization capacity, reinforcing the statistical consistency of the results.

**Table 7:**
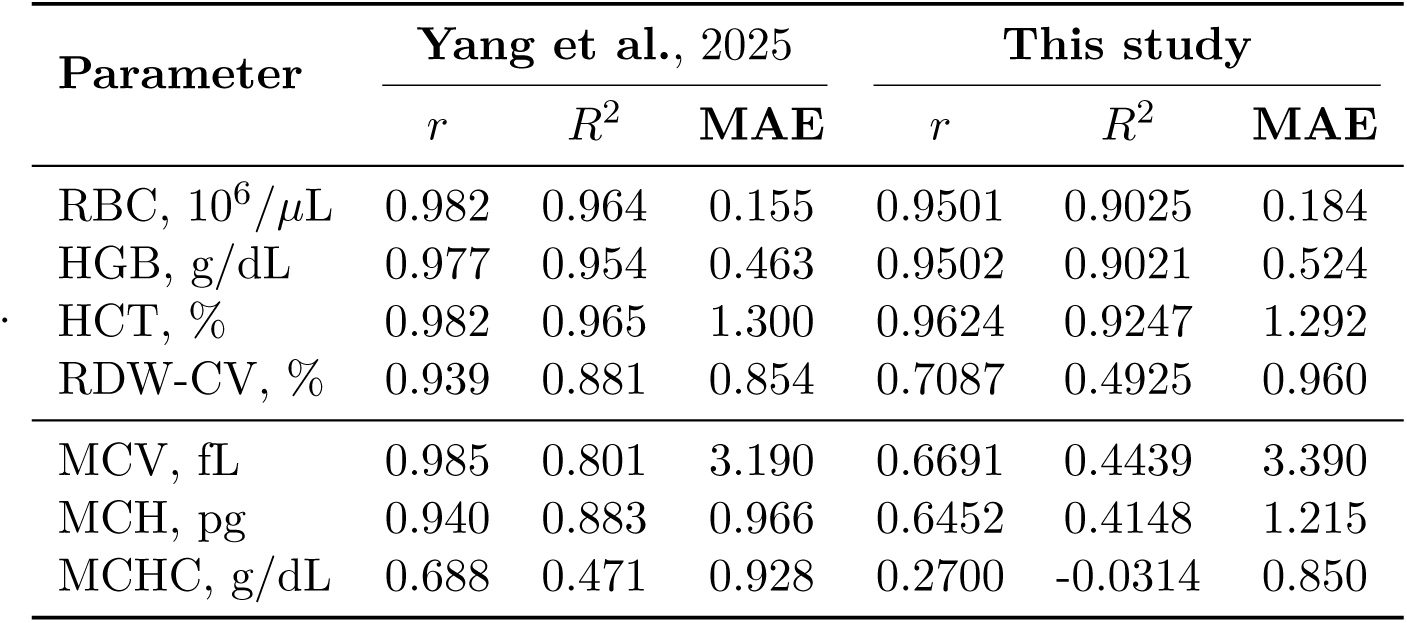
Performance comparison between Yang et al. [. **6] and the present study**

| Parameter | Yang et al., 2025 |  |  | This study |  |  |
| --- | --- | --- | --- | --- | --- | --- |
| | $r$ | $R^2$ | MAE | $r$ | $R^2$ | MAE |
| RBC, $10^6/\mu\text{L}$ | 0.982 | 0.964 | 0.155 | 0.9501 | 0.9025 | 0.184 |
| HGB, g/dL | 0.977 | 0.954 | 0.463 | 0.9502 | 0.9021 | 0.524 |
| HCT, % | 0.982 | 0.965 | 1.300 | 0.9624 | 0.9247 | 1.292 |
| RDW-CV, % | 0.939 | 0.881 | 0.854 | 0.7087 | 0.4925 | 0.960 |
| MCV, fL | 0.985 | 0.801 | 3.190 | 0.6691 | 0.4439 | 3.390 |
| MCH, pg | 0.940 | 0.883 | 0.966 | 0.6452 | 0.4148 | 1.215 |
| MCHC, g/dL | 0.688 | 0.471 | 0.928 | 0.2700 | -0.0314 | 0.850 |

According to Yang et al. [6], the MAE value obtained for hemoglobin was considered clinically acceptable because it was below the standard deviation reported by the International Council for Standardization in Hematology, ICSH, for internal quality control. Under the same criterion, the MAE value for hemoglobin observed in the present study would also be considered acceptable as proposed by the authors.

It is important to note that indicators of analytical agreement such as MAE, *R*^2^, *r*, and Bland–Altman analysis are necessary but not sufficient to conclude clinical feasibility, because these metrics predominantly describe average performance and overall agreement and do not fully characterize the magnitude and distribution of individual errors, which can be clinically relevant in specific cases.

Thus, even in multicenter studies such as that of Yang et al. [6], the clinical consolidation stage of the method requires comparing performance against explicit acceptability criteria such as those from the Clinical Laboratory Improvement Amendments, CLIA, and/or specifications based on within-subject biological variation and internal analytical system variation limits.

Finally, it is important to emphasize that both studies were developed independently, without prior knowledge of each other during their respective execution, which reinforces the convergence of results and supports the robustness of the proposed methodological approach.

## 5 Limitations

This study has limitations that should guide future work. The small sample size and single-center data collection limit the generalizability of the findings, indicating the need for multicenter validation. The presence of outliers in the Bland–Altman analysis suggests that the method may produce clinically relevant errors in specific cases and is currently more suitable for screening than for definitive diagnosis.

The use of a single microscope, combined with variability in capture position and magnification level of the images, may have limited extraction of more subtle morphological features, especially for variables dependent on cellular heterogeneity such as RDW-CV. Strategies based on higher magnification or individual erythrocyte segmentation could provide more discriminative information and improve predictive performance.

In addition, traditional smear preparation can introduce variations in cell dispersion and overlap, indicating that more homogeneous preparation techniques represent an opportunity for further improvement. Direct prediction of derived indices such as MCV, MCH, and MCHC could reduce error propagation associated with sequential calculation. Finally, this work evaluated technical feasibility and analytical agreement of the method and was not designed for formal clinical or regulatory validation at this stage.

## 6 Conclusion

This study demonstrated that artificial intelligence models can estimate with high accuracy the main erythrocyte indices RBC, HGB, and HCT from blood smear images obtained directly through the eyepiece of conventional optical microscopes. The models achieved *R*^2^ values above 0.90 for RBC, HGB, and HCT, and moderate performance for RDW-CV with *R*^2^ = 0.49. These results indicate high agreement with reference laboratory measurements, with MAE of 0.184 for RBC, 0.524 for HGB, 1.292 for HCT, and 0.960 for RDW-CV.

The proposed methodology reduces dependence on automated scanners and high-cost hematology analyzers, enabling its application in small laboratories, public health programs, and remote communities. This characteristic gives the method strong potential impact in democratizing access to health care, including in settings with limited infrastructure.

Future research should prioritize multicenter validation and dataset expansion, as well as investigation of alternative slide preparation methodologies that promote greater homogeneity in cell distribution regardless of the analyzed field. Additionally, strategies aimed at more detailed extraction of morphological information, such as use of higher magnification or individual cell segmentation, may contribute to improved predictive performance. As a subsequent step, consolidation of the method requires evaluation in light of regulatory criteria such as those defined by CLIA, as well as comparison with internal analytical system variation limits and specifications based on intra-biological variation.

## Acknowledgments

The author thanks Laboratório Hipólito Monte for the partnership and HORIBA do Brasil for providing the HEMAPREP device.

## Author Contributions

LP: Conceptualization, methodology, data analysis, manuscript writing.

## Data and Supplementary Material Availability

All data, datasets, standardized experimental protocols, and trained model checkpoints generated in this study are publicly available at https://github.com/lspraciano/erythrogram-supply-data

